# New frontiers in health literacy: Using ChatGPT to simplify health information for people in the community

**DOI:** 10.1101/2023.07.24.23292591

**Authors:** Julie Ayre, Olivia Mac, Kirsten McCaffery, Brad R McKay, Mingyi Liu, Yi Shi, Atria Rezwan, Adam G Dunn

**Author notes:** Corresponding author: Julie Ayre, Rm 128C Edward Ford Building, The University of Sydney, NSW.

## Abstract

**Background:** Most health information does not meet the health literacy needs of our communities. Writing health information in plain language is time-consuming but the release of tools like ChatGPT may make it easier to produce reliable plain language health information.

**Objective:** To investigate the capacity for ChatGPT to produce plain language versions of health texts.

**Design:** Observational study.

**Subjects:** Twenty-six health texts from reputable websites.

**Interventions:** ChatGPT was prompted to ‘rewrite the text for people with low literacy.’ Researchers captured three revised versions of each original text.

**Main Measures:** Objective health literacy assessment, including Simple Measure of Gobbledygook (SMOG), proportion of the text that contains complex language (%), number of instances of passive voice, and subjective ratings of key messages retained (%).

**Key Results:** On average, original texts were written at Grade 12.8 (SD=2.2) and revised to Grade 11.0 (SD=1.2), p<0.001. Original texts were on average 22.8% complex (SD=7.5%) compared to 14.4% (SD=5.6%) in revised texts. Original texts had on average 4.7 instances (SD=3.2) of passive text compared to 1.7 (SD=1.2) in revised texts. On average 80% of key messages were retained. The more complex original texts showed more improvements than less complex original texts. For example, when original texts were ≥ Grade 13, revised versions improved by an average 3.3 Grades (SD=2.2), p<0.001. Simpler original texts (< Grade 11) improved by an average 0.5 grades (SD=1.4), p<0.001.

**Conclusions:** This study used multiple objective assessments of health literacy to demonstrate that ChatGPT can simplify health information while retaining most key messages. However, the revised texts typically did not meet health literacy targets for grade reading level, and improvements were marginal for texts that were already relatively simple.

---

In the wake of COVID-19, health literacy has come to the forefront of public health research and practice, with persistent calls to provide health information that is easy to access and understand (1, 2). COVID-19 information has been continually assessed as too complex for people to understand, particularly for groups that may have low health literacy, such as those who are older, with lower education, and who have less fluency in a community’s dominant language (3–6). This issue is not limited to COVID-19. Studies consistently report that most health information does not address the health literacy needs of our communities. This includes information developed by government, health services, and non-government organisations (7, 8).

Addressing this issue is challenging given the vast amount of health information available online. Currently, writing in plain language requires a health information provider to manually implement advice from health literacy guidelines and checklists (9–12). This is a process that demands considerable expertise and time. Though there are tools for objectively assessing the health literacy of health information and automating text-simplification (13–15), revisions are still largely carried out by humans.

Recent advances in large language models present new opportunities that might transform our ability to develop plain language health information at scale. For example, in November 2022, OpenAI publicly released ChatGPT, a large language model that has been trained on a large database of text data to produce plausible, contextually appropriate, and human-like responses to *prompts*—typically questions or requests to produce writing meeting certain constraints. Large language models do not synthesise or evaluate evidence, but rather they predict what should come next in a piece of text by learning from large volumes of training data (16). ChatGPT is also capable of adapting text to different writing styles and audiences, has a simple user interface that does not require software or programming expertise, and is freely available.

There is limited evidence showing that ChatGPT can produce information that adheres to health literacy guidelines. Ali et al. (17) prompted ChatGPT to write patient-facing letters suitable for an 11–12-year-old, a target that aligns with health literacy recommendations in the US. On average, ChatGPT produced output at a US 9^th^ grade reading level (14–15 years), well above the prompt instructions. Ayoub et al. (18) used a well-established subjective health literacy instrument, the Patient Education Materials Assessment Tool, to evaluate the quality of postoperative patient instructions generated by ChatGPT. These instructions were rated as adequately understandable, actionable and generally complete (i.e. the instructions included 75% to 100% of key steps). However only 8 texts were generated and the grade reading levels of the texts were not reported, despite this being explicit in the ChatGPT prompt. Whilst these studies provide some insight into using ChatGPT to create plain language information, there is substantial room for improvement, both in terms of optimising the ChatGPT prompts and employing more comprehensive assessment of plain language. Other studies looking at ChatGPT outputs in health domains have found that e.g. simplified summaries of radiology reports were generally correct and complete, with low potential for harm, though the complexity of the language was not assessed (19, 20). Several studies have also identified a reasonable level of accuracy in ChatGPT output that responds to health questions (21–24).

This study sought to investigate the capacity for ChatGPT (GPT-3.5) to produce plain language versions of health texts across a range of health topics. To our knowledge no studies have evaluated the appropriateness of plain language health information generated by ChatGPT using multiple objective assessments.

## Methods

### Text selection

The research team collected extracts from online health information published by recognised national and international health information provider websites such as the World Health Organization, Centers for Disease Control and Prevention, and National Health Service (UK) (Appendix A). Extracts were at least 300 words and did not rely on images to explain the text.

### ChatGPT

ChatGPT-3.5 was accessed via chat.openai.com between 28 April 2023 and 8 May 2023. The platform allows users to ‘converse’ with the model via API, by sending text-based prompts which the model then responds to. The model seeks to supply users with plausible, human-like responses. However, responses reflect statistical patterns based on training data, rather than knowledge synthesis (16). Given the risks associated with delivering unsupervised health advice, ChatGPT includes some safeguards to reduce unsafe or harmful prompts. For example, the model is known not to give personalised health advice.

### ChatGPT prompt development and text revision

To develop a prompt that applies health literacy principles to written text, several prompts were tested on four sample texts. Two types of prompts were tested: (a) prompts that described specific health literacy principles (e.g. simple language, active voice, minimal jargon); and (b) prompts that described the target audience. The latter reflected typical health literacy priority groups such as people who do not speak English as their main language, people who read at a school student level, people without health or medical training (25).

Each candidate prompt was used in a separate ‘chat’ to reduce the risk of interference from previous instructions to revise other texts (13 March to 11 April 2023). The research team generated two revised texts per candidate prompt and assessed these for grade reading score, complex language, passive voice, and subjective appraisals of retention of key messages (Appendix B). Findings were discussed across the whole research team. The prompt ‘*rewrite the text for people with low literacy*’ was ultimately selected for this study because it more consistently produced texts with a lower grade reading score across the four sample texts and each of two iterations, avoided passive voice, used simpler language, and is a brief prompt that is easy to use. We collected three responses from each prompt using the ‘regenerate’ function.

### Text assessment

Each text was assessed using the Sydney Health Literacy Lab Health Literacy Editor, which we developed (15). This is an online tool designed to objectively assess the extent that written health information is written in plain language. Four assessments were obtained: number of words, grade reading score, complex language and passive voice (Table 1).

**Table 1.** Objective assessments of text health literacy.

| Assessment | Description and rationale |
| --- | --- |
| Number of words | Number of words is not a health literacy assessment but provides context about the extent that ChatGPT ‘summarises’ the text. |
| Grade reading score | Grade reading score estimates how difficult a text is to read, and roughly corresponds to US school grade reading levels. It is an assessment identified in many health literacy guidelines, and widely used in health literacy research. In Australia a Grade reading score of 8 or lower is a common target (see for example, Clinical Excellence Commission (26)).<br>The Editor assesses grade reading score using the Simple Measure of Gobbledygook (SMOG) (27). The Editor provides accurate assessment of |
|  | SMOG when compared to the reference standard (hand-calculations) and other software (28). |
| **Complex language** | Health literacy guidelines consistently recommend using common, everyday language (9-12). The Editor's 'complex language' assessment estimates the extent that a text adheres to this principle.<br>This is presented as the proportion of the text (%) that contains acronyms, uncommon words (as defined by an existing English-language corpus), or terms listed in the Editor's simple English thesaurus. Although there is no established 'cut-off' or 'target' level of complex language, lower scores indicate lower levels of complex language. For each text, the research team identified up to 5 key topic words that were excluded from complex language assessment as these words were inherent to the text. |
| **Passive voice** | Many health literacy guidelines recommend writing in the active voice, advising to avoid more than a single instance of passive voice in a given text (9, 10). The Editor assesses the number of instances of passive voice using natural language processing to parse the text for this grammatical construction. |
| **Dot points for lists** | Using dot points for long lists is recommended in some plain language guidelines (11, 29). |

Completeness was assessed by subjectively rating whether the key messages were retained in each text. Key messages were developed independently by authors JA and OM, with discrepancies resolved through discussion. The two people who assessed the completeness of the revised text were not involved in selecting the text or developing key messages. One consumer and one academic researcher rated each text (84.3% agreement across 510 message ratings). Scores represent the average number of key messages retained across both assessors.

### Analysis

Descriptive statistics were calculated for each text and averaged across the three texts generated by the ChatGPT prompt. Results also present the minimum and maximum scores of individual ChatGPT revisions to provide a sense of the reliability of the prompt. Differences between original and revised text assessments were analysed using paired-sample t tests. ANOVA was used to explore these differences across texts with low, medium and high complexity in the original versions, and Pearson’s correlations was used to explore the relationships between continuous variables.

## Results

On average the 26 original texts had a grade 12.8 reading level. Almost one quarter (22.8%) of the words were assessed as ‘complex’ and there were on average 4–5 instances of passive voice (Table 2). Texts revised by ChatGPT were on average 1.8 grade reading scores lower (M=11.0, p<0.001), with significantly less complex language (14.4%, p<0.001) and less use of passive voice (1.7, p<0.001). Fourteen of the 26 original texts (54%) showed lists as dot points. When these texts were revised, only 4 of the 56 revised versions (7%) used the same format. No revised texts introduced dot points where there were none in the original text.

**Table 2.** Summary of objective text characteristics, original and revised texts (N=26)

| Assessment | Original text | Average ChatGPT revised text |  |  | Decrease |
| --- | --- | --- | --- | --- | --- |
|  | M (SD) | M (SD) | Min | Max | M (SD) |
| Number of words | 420.2 (89.3) | 228.9<br>(51.1) | 88 | 462 | 191.2 (19.7) |
| Grade reading score | 12.8 (2.2) | 11.0 (1.2) | 8.3 | 14.5 | 1.8 (0.4) |
| Complex language (% of the text) | 22.8 (7.5) | 14.4 (5.6) | 3.2 | 37.8 | 8.4 (1.1) |
| Passive voice (n) | 4.7 (3.2) | 1.7 (1.2) | 0 | 6 | 3 (0.5) |

*Notes:* Minimum and maximum scores represent the lowest and highest scores recorded for any ChatGPT text. Target for grade reading score is Grade 8, there is no target for complex language (but lower scores are more favourable), target for passive voice is <2 instances. \*All differences $p<0.001$

ChatGPT was also more effective at revising texts that were more complex to begin with (Table 3). For example, when ChatGPT revised texts that were originally grade 13 or higher, the grade reading score was lowered by an average 3.3 grades. This was a much larger improvement than revisions to texts that were originally Grade 11 or lower (mean decrease of 0.5, p=0.009) or that were originally Grades 11 to 12 (mean decrease of 1.4, p=0.032). Similar patterns were observed for complex language and passive voice.

**Table 3.**
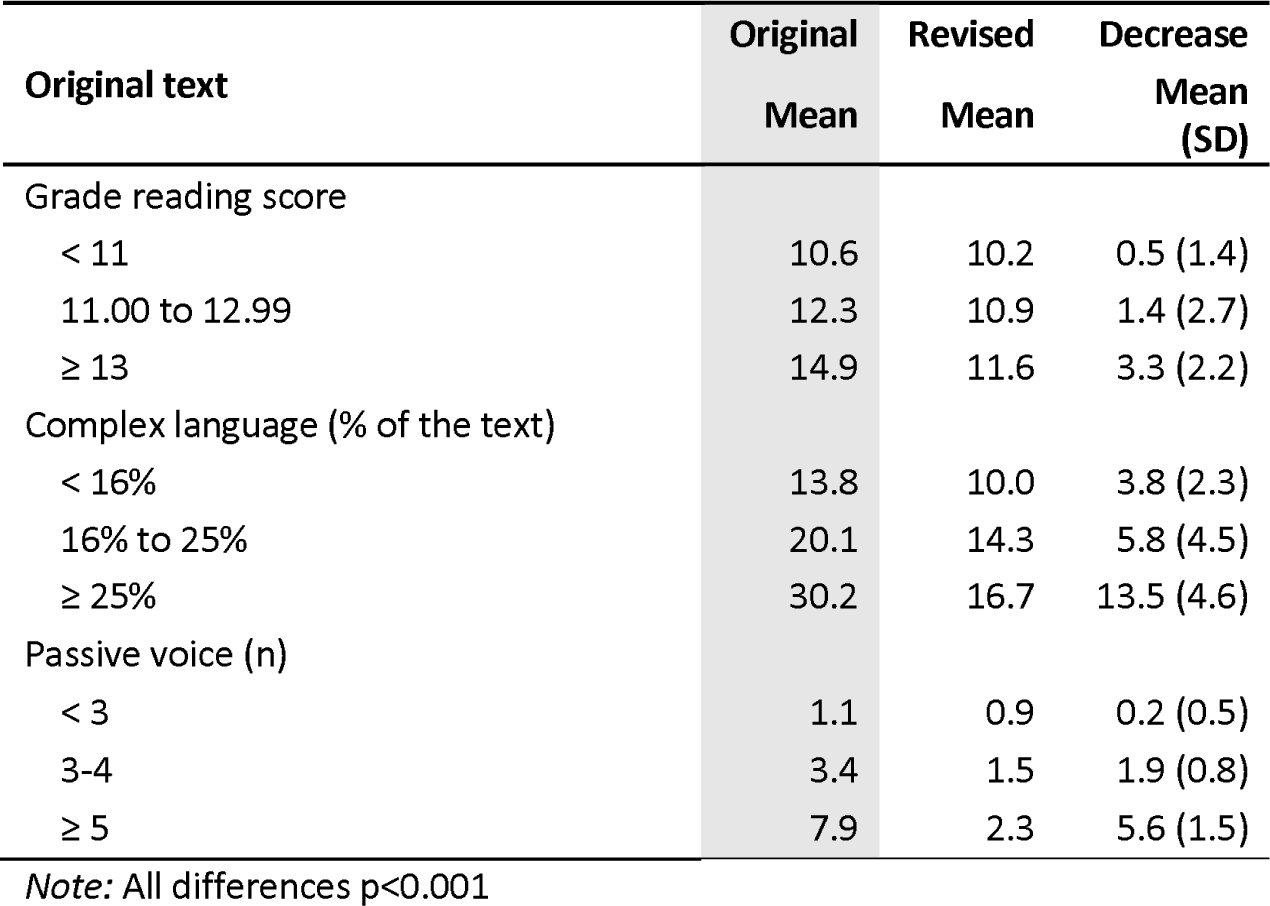
Summary of ChatGPT improvements, by original text complexity (N=26)

| Original text | Original<br>Mean | Revised<br>Mean | Decrease<br>Mean<br>(SD) |
| --- | --- | --- | --- |
| Grade reading score |  |  |  |
| < 11 | 10.6 | 10.2 | 0.5 (1.4) |
| 11.00 to 12.99 | 12.3 | 10.9 | 1.4 (2.7) |
| ≥ 13 | 14.9 | 11.6 | 3.3 (2.2) |
| Complex language (% of the text) |  |  |  |
| < 16% | 13.8 | 10.0 | 3.8 (2.3) |
| 16% to 25% | 20.1 | 14.3 | 5.8 (4.5) |
| ≥ 25% | 30.2 | 16.7 | 13.5 (4.6) |
| Passive voice (n) |  |  |  |
| < 3 | 1.1 | 0.9 | 0.2 (0.5) |
| 3-4 | 3.4 | 1.5 | 1.9 (0.8) |
| ≥ 5 | 7.9 | 2.3 | 5.6 (1.5) |

Note: All differences $p < 0.001$

Original texts had on average 6.5 key messages (SD=2.0), with a range of 3 to 10. On average 79.8% of key messages were retained in revised texts (SD=15.0), ranging from 20% in one instance to as high as 100%. Completeness of revised texts was not related to the number of key messages in an original text (p=0.43), its length (p=0.84), or health literacy assessment (grade reading score: p=0.39; complex language: p=0.53; passive voice: p=0.68).

## Discussion

When asked to simplify existing health information, ChatGPT on average improved the grade reading score of texts, used less complex language, and removed instances of the passive voice. It achieved this while retaining 80% of the key messages. These improvements were particularly notable for texts that were more complex to begin with. For example, ChatGPT reduced the grade reading score for the most complex texts by an average 3 grade reading levels (original: 14.9 vs revised: 11.6). While there is no clear reference for what makes a meaningful difference to patients, we found that a change of 3 grade reading levels is still substantial even though it is above the target of a Grade 8 reading level. By comparison, when original texts were already below Grade 11, the revised versions were only half a grade lower, suggesting ChatGPT may be less useful for simpler texts. Together this suggests that ChatGPT may provide a useful ‘first draft’ of plain language health information that can be further refined through human revision and checking processes.

These findings are consistent with other studies evaluating the capacity of ChatGPT to develop community-facing health information. For example, clinicians have rated ChatGPT summaries of radiology reports as relatively accurate, clear and concise (19, 20). A previous study also reported that ChatGPT typically produced health information above a grade 8 reading level (17). However, the prompt used in the current study generated texts of a lower grade reading score than the previous study, which produced a SMOG grade reading score of 12.5 (17) compared to our score of 11.0.

These findings highlight some of the benefits and limitations of using ChatGPT to improve access to plain language health information. Several studies now report that the platform generates relatively accurate health information and can adequately retain key messages when revising texts, although human and clinical oversight is needed to ensure that text is correct, all key messages are retained, and phrasing is coherent and natural (17–22). Due to ChatGPT’s reliance on human input for training, users should also carefully reflect on its potential to perpetuate biases relating to e.g. race, age, gender, and ethnicity (16). The current study also demonstrated that ChatGPT can support implementation of health literacy guidelines for written health information (9–12). Although it is not a complete solution, ChatGPT’s strength lies in the speed at which it can redraft plain language content for further review, rather than its ability to generate a ‘final’ public-facing resource.

This study had several strengths. We evaluated the use of ChatGPT across a wide range of health topics, generated three versions of each text, and used multiple objective health literacy assessments. Key messages were developed prior to the study and key message retention ratings were double coded, including by a consumer. Lastly, by documenting how the prompt was developed we highlight the potential pitfalls of other prompts to our readers.

The main limitation of this study is that we did not evaluate how easily consumers could understand the revised texts, either using subjective assessment such as Likert rating scales or objective assessment such as knowledge questions. Other limitations are that ChatGPT will continue to evolve and will likely improve over time. Results presented in this study reflect ChatGPT-3.5, at the time of data collection, and do not reflect the performance of more recent versions of ChatGPT, which may become more widely used in the future.

Future research could vary the parameters of the original texts. For example, it is unclear how well ChatGPT can simplify information for less prevalent health conditions, different types of resources, longer texts, and texts written in different languages or for different cultural contexts. Research could also explore other emerging publicly accessible interfaces to large language models such as Google Bard and Bing Chat. In this study, no personal information is included in the original text because the information is general, but in cases where personal information about a diagnosis or prognosis is used in bespoke patient information, additional issues related to data privacy and ethical concerns may become an issue. With further evidence that ChatGPT can reliably, ethically, and safely produce health information that most people can easily understand, it would be valuable to explore how the platform can be systematically implemented into health literacy tools and health organisation practices.

Interfaces into large language models have the potential to rapidly transform the way plain language health information is produced, especially given the rapid improvements to large language models and the interfaces that make them accessible and useful. This study used multiple objective assessments of health literacy to demonstrate that ChatGPT was able to simplify health information while retaining key messages. However, human oversight remains essential to ensure accuracy, completeness, and effective application of health literacy guidelines. Further research is needed to identify how health information providers can best leverage this technology to meet the health literacy needs of our communities.

## Data Availability

All data produced are available in Appendix

## Contributors

We would like to acknowledge the contributions of our consumer partners on this project: Atria Rezwan, Lauren Resnick, Peta de-Haan, Debra Letica and Oliver Slewa.

## Funders

Dr Ayre is supported by a National Health and Medical Research Council fellowship (APP 2017278).

## Prior presentations

None.

## Conflicts of interest

Members of the research team (JA, KM) are directors of a health literacy consultancy (Health Literacy Solutions Ltd., Pty). No other declared conflicts of interest.

## Appendix A

Excel data file

## Appendix B

### ChatGPT prompt development

#### Prompt objective

To rewrite health information in plain language, in alignment with health literacy recommendations.

#### Health literacy principles of interest

Principles are taken from the Universal Precautions Toolkit and Patient Education Materials Assessment Tool. These include:

1. Text uses plain language (use common, everyday language; avoid jargon; use the active voice where possible). Assessed objectively using the Sydney Health Literacy Lab (SHeLL) Health Literacy Editor.

- Grade reading score: Target Grade 8.0 or lower; Adequate: Grade 10.0
- Complex language: Target <15%; Adequate <20% (based on author expertise)
- Passive voice: Target: 0-1^2^, Adequate: 2
2. Text directly addresses the reader
3. Text defines medical terms and acronyms the first time they are used.

Each prompt was testing in a new chat window within ChatGPT. JA and OM generated two revised versions across 4 different texts (sciatica, dementia, leukaemia, malaria) for each prompt that was trialled. Prompts were also assessed in terms of retention of key messages, level of detail, accuracy, and introduction of new information.

### Observations - prompt categories

#### References to age and school grade

- Referring to an age bracket rather than a school grade typically returned a lower grade reading score
- This also reduced complex language and typically removed all passive voice
- Prompting for 11 years old produced more natural language/better responses than for younger e.g. 10 years old and maintained most of the detail.
- Sometimes the revised text would reference age e.g. “Tell an adult” rather than “See your doctor” and “people like you” when referring to children aged <14 years.
- Issues with stigma in this phrasing

#### People with English as a second language / non-native English speakers

- Referring to non-native English speakers (in itself) did not tend to reduce grade reading score, complex language, or passive voice. When combined with reference to age/school, these scores improved (grade reading score ∼8-11, low complex language and low/zero passive voice).

#### People with low literacy

- Similar to the combination of age/school + English as a second language
- Typically retained detail and language read well, tone of original text was retained.
- Occasionally text was addressing a child

#### Explicit instructions describing health literacy principles

- Grade reading score tended to be high
- Passive voice often not removed, even when this was an explicit instruction. Sometimes ‘active’ voice was interpreted by ChatGPT as being more conversational / informal

#### Simplified language

- Grade reading score and complex language tended to be lower, with passive voice removed
- Revisions retained detail and key messages

### Prompt shortlist

#### Prompt shortlist comprised the following

1. Rewrite the text into simplified language that is easy to understand
2. *Rewrite the text so it is suitable for non-native English speakers and people with low literacy*
3. *Rewrite the text for people with low literacy*

#### Objective assessments of shortlist prompts

JA and OM generated two revised versions across 4 different texts (sciatica, dementia, leukaemia, malaria) for each prompt that was trialled.

The tables below present the objective scores for grade reading score, complex language and passive voice. Coloured cells show the extent that the texts generated by ChatGPT met target health literacy recommendations. Cells that are green indicate at least one of the two texts had a score in the target range. Cells that are yellow indicate at least one of the two texts had a score in the adequate range. Option 3 ‘*Rewrite the text for people with low literacy’* was selected for use in this study.

#### Grade reading score

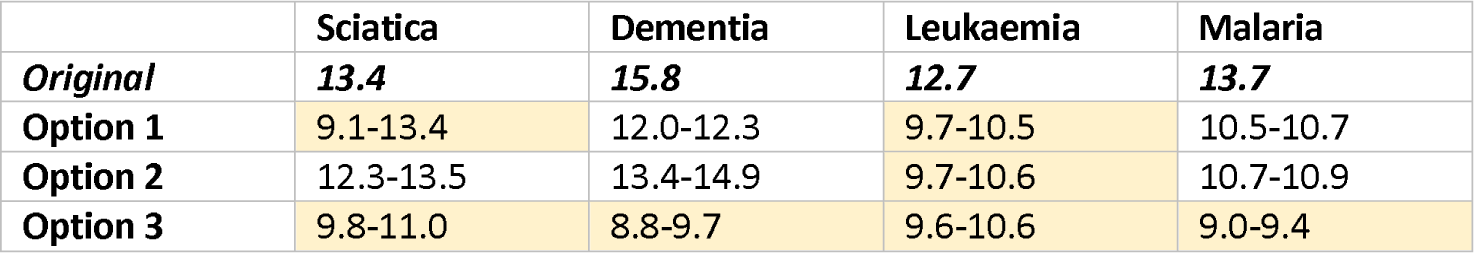

#### Complex language

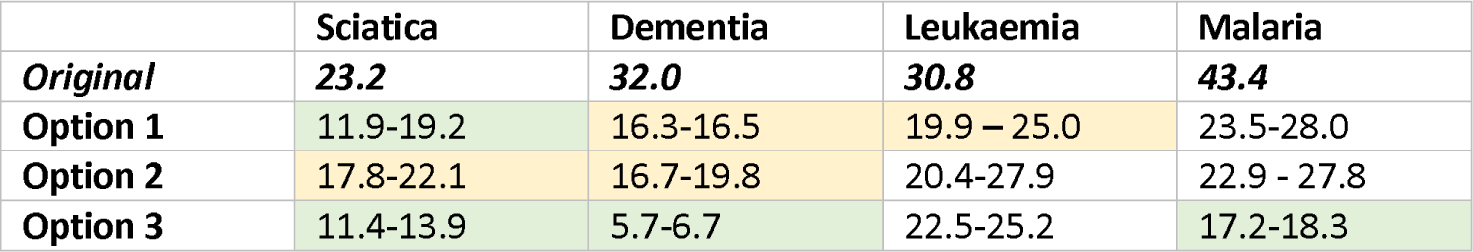

#### Passive voice

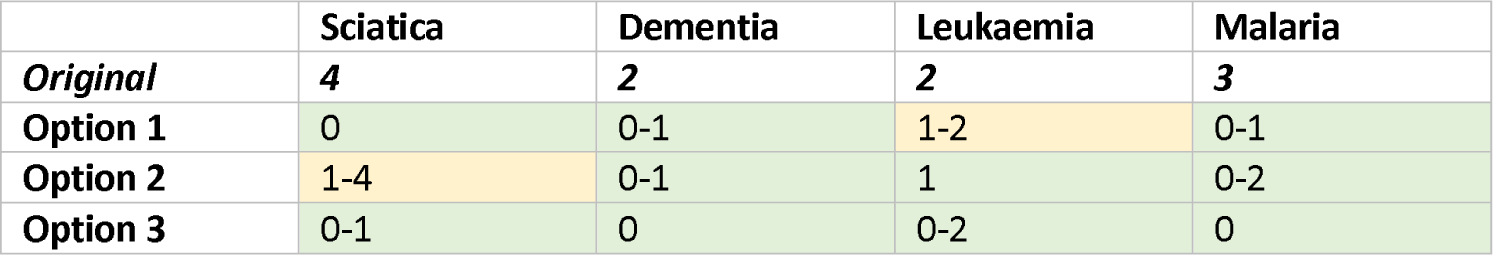

