## Appendix A and B for "New frontiers in health literacy: Using ChatGPT to simplify health information for people in the community": Appendix B ChatGPT prompt development.docx

**Grade reading score**

|  | Sciatica | Dementia | Leukaemia | Malaria |
| --- | --- | --- | --- | --- |
| *Original* | ***13.4*** | ***15.8*** | ***12.7*** | ***13.7*** |
| Option 1 | 9.1-13.4 | 12.0-12.3 | 9.7-10.5 | 10.5-10.7 |
| Option 2 | 12.3-13.5 | 13.4-14.9 | 9.7-10.6 | 10.7-10.9 |
| Option 3 | 9.8-11.0 | 8.8-9.7 | 9.6-10.6 | 9.0-9.4 |

**Complex language**

|  | Sciatica | Dementia | Leukaemia | Malaria |
| --- | --- | --- | --- | --- |
| *Original* | ***23.2*** | ***32.0*** | ***30.8*** | ***43.4*** |
| Option 1 | 11.9-19.2 | 16.3-16.5 | 19.9 **–** 25.0 | 23.5-28.0 |
| Option 2 | 17.8-22.1 | 16.7-19.8 | 20.4-27.9 | 22.9 - 27.8 |
| Option 3 | 11.4-13.9 | 5.7-6.7 | 22.5-25.2 | 17.2-18.3 |

**Passive voice**

|  | Sciatica | Dementia | Leukaemia | Malaria |
| --- | --- | --- | --- | --- |
| *Original* | ***4*** | ***2*** | ***2*** | ***3*** |
| Option 1 | 0 | 0-1 | 1-2 | 0-1 |
| Option 2 | 1-4 | 0-1 | 1 | 0-2 |
| Option 3 | 0-1 | 0 | 0-2 | 0 |
